# Maximizing the impact of limited vaccine supply under different epidemic conditions: a two-city monkeypox modelling analysis

**DOI:** 10.1101/2022.08.18.22278949

**Authors:** Jesse Knight, Darrell H.S. Tan, Sharmistha Mishra

## Abstract

**background:** In the current global monkeypox outbreak, many jurisdictions have been faced with limited vaccine supply, motivating interest in efficient allocation. We sought to explore optimal vaccine allocation between two linked transmission networks over a short-term time horizon, across a range of epidemic conditions.

**methods:** We constructed a deterministic compartmental sveir model of monkeypox transmission. We parameterized the model to reflect two representative, weakly connected gbmsm sexual networks (cities) in Ontario. We simulated roll-out of 5000 vaccine doses over 15 days, starting 60 days after epidemic seeding with 10 imported cases. Within this model, we varied: the relative city (network) sizes, epidemic potentials (*R*_0_), between-city mixing, and distribution of imported/seed cases between cities. In each context (combination of varied factors), we then identified the “optimal” allocation of doses between cities — resulting in the fewest cumulative infections by day 120.

**results:** Under our modelling assumptions, we found that a fixed supply of vaccines could generally avert more infections over short-term time horizons when prioritized to: a larger transmission network, a network with more initial infections, and/or a network with greater *R*_0_. Greater between-city mixing decreased the influence of initial seed cases, and increased the influence of city *R*_0_ on optimal allocation. Under mixed conditions (e.g. fewer seed cases but greater *R*_0_), optimal allocation saw doses shared between cities, suggesting that proximity-based and risk-based vaccine prioritization can work in combination to minimize transmission.

**interpretation:** Prioritization of limited vaccine supply based on network-level risk factors can help minimize transmission during an emerging epidemic. Such prioritization should be grounded in an understanding of context-specific drivers of risk, and should acknowledge the potential connectedness of multiple transmission networks.

## 1 Introduction

The emerging outbreak of monkeypox virus (mpvx) worldwide includes 1,112 cases in Canada as of 2022 August 17 [1]. A third-generation replication-deficient smallpox vaccine (Imvamune^®^) has been licensed for use against monkeypox and related orthopoxviruses in Canada since 2020, for the purpose of national security [2]. Shortly after local cases were reported, rapid pre-exposure prophylaxis vaccination efforts were initiated to help reduce acquisition, infectivity, and/or disease severity among communities disproportionately affected by mpvx, including gay, bisexual, and other men who have sex with men (gbmsm) [3]. However, many jurisdictions, across countries and within Canada, were faced with a limited local supply of vaccines during the first few weeks of mpvx outbreak.

It is well-established that prioritizing a limited supply of vaccines to sub-populations experiencing disproportionately higher risk — individual-level and/or network-level acquisition and/or transmission risk — can maximize infections averted [4,5]. Such networks may have different characteristics that shape the epidemic potential (*R*_0_) within the network itself. A network’s connectedness to other networks further shapes the chances and number of imported cases by the time vaccine allocation decisions and roll-out begin.

We sought to explore optimal allocation of a fixed supply of mpvx vaccine across two jurisdictions — i.e. weakly connected transmission networks — under different epidemic conditions. Specifically, we explored differences between two jurisdictions in: population size of gbmsm; epidemic potential (*R*_0_); imported/seed cases; and connectedness of the two jurisdictions. The goal of this modeling study was to produce fundamental and generalizable insights into mpvx vaccine prioritization in the context of interconnected sexual networks, using jurisdictions (cities) within a province like Ontario, Canada as an example.

## 2 Methods

We constructed a deterministic compartmental sveir (susceptible, vaccinated, exposed, infectious, recovered) model of mpvx transmission. The modelled population aimed to represent the Ontario gbmsm community, and included two levels of sexual risk (higher, lower) and two weakly connected transmission networks (cities A, B). Figure 1a illustrates the modelled city/risk strata, Figure 1b illustrates the sveir health states, and Table 1 summarizes the default model parameters. To parameterize the model, we drew on prior analyses of gbmsm sexual networks in Canada [6,7], and emerging mpvx epidemiological data in the context of the current epidemic [8–12]. Appendix A provides additional details about the model implementation and parameterization.

**Table 1:**
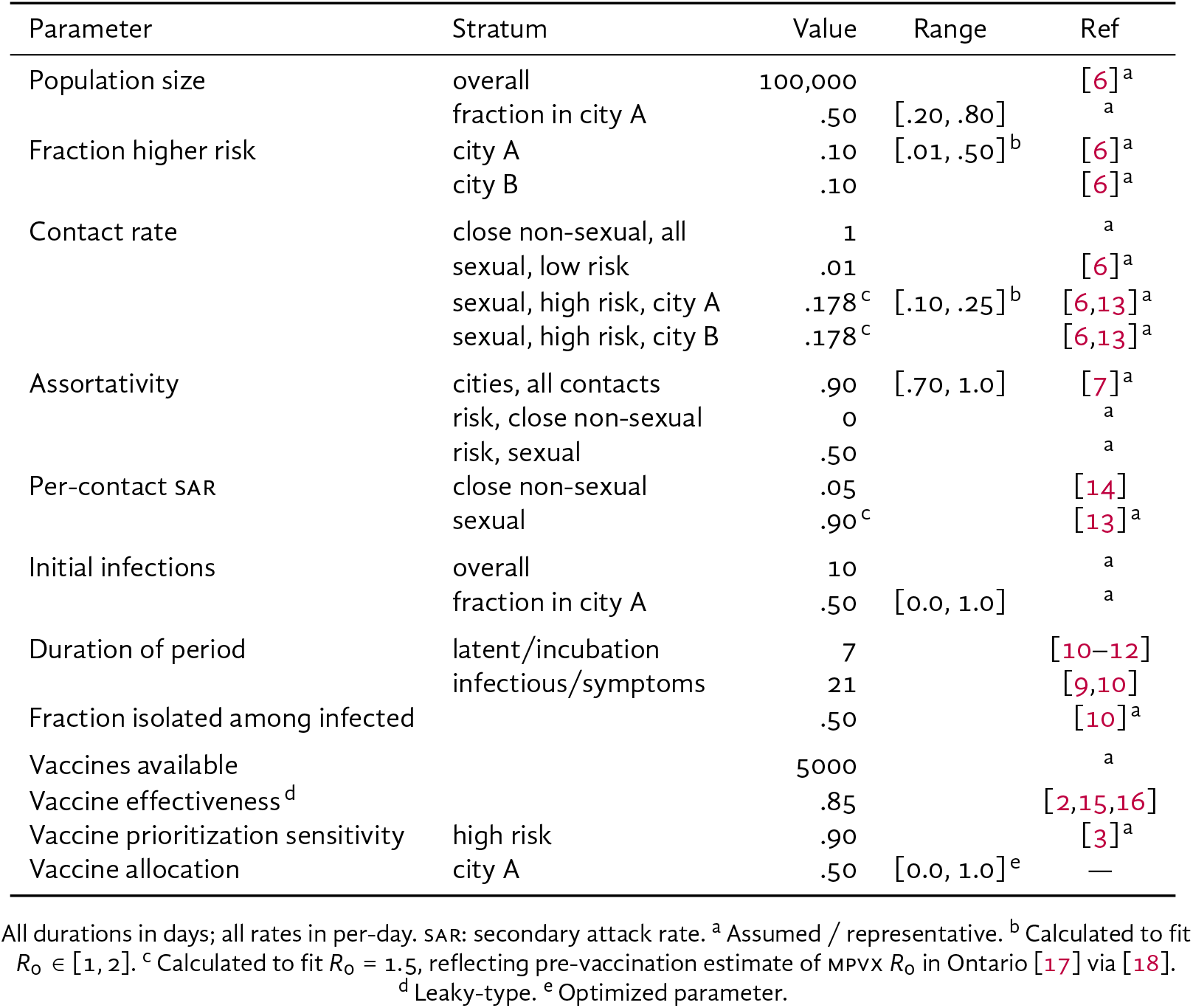
Model parameters, including default values and ranges explored via grid sweep

| Parameter | Stratum | Value | Range | Ref |
| --- | --- | --- | --- | --- |
| Population size | overall | 100,000 |  | [6]<sup>a</sup> |
|  | fraction in city A | .50 | [.20, .80] | <sup>a</sup> |
| Fraction higher risk | city A | .10 | [.01, .50]<sup>b</sup> | [6]<sup>a</sup> |
|  | city B | .10 |  | [6]<sup>a</sup> |
| Contact rate | close non-sexual, all | 1 |  | <sup>a</sup> |
|  | sexual, low risk | .01 |  | [6]<sup>a</sup> |
|  | sexual, high risk, city A | .178<sup>c</sup> | [.10, .25]<sup>b</sup> | [6,13]<sup>a</sup> |
|  | sexual, high risk, city B | .178<sup>c</sup> |  | [6,13]<sup>a</sup> |
| Assortativity | cities, all contacts | .90 | [.70, 1.0] | [7]<sup>a</sup> |
|  | risk, close non-sexual | 0 |  | <sup>a</sup> |
|  | risk, sexual | .50 |  | <sup>a</sup> |
| Per-contact SAR | close non-sexual | .05 |  | [14] |
|  | sexual | .90<sup>c</sup> |  | [13]<sup>a</sup> |
| Initial infections | overall | 10 |  | <sup>a</sup> |
|  | fraction in city A | .50 | [0.0, 1.0] | <sup>a</sup> |
| Duration of period | latent/incubation | 7 |  | [10–12] |
|  | infectious/symptoms | 21 |  | [9,10] |
| Fraction isolated among infected |  | .50 |  | [10]<sup>a</sup> |
| Vaccines available |  | 5000 |  | <sup>a</sup> |
| Vaccine effectiveness<sup>d</sup> |  | .85 |  | [2,15,16] |
| Vaccine prioritization sensitivity | high risk | .90 |  | [3]<sup>a</sup> |
| Vaccine allocation | city A | .50 | [0.0, 1.0]<sup>e</sup> | — |

All durations in days; all rates in per-day. SAR: secondary attack rate. <sup>a</sup> Assumed / representative. <sup>b</sup> Calculated to fit $R_0 \in [1, 2]$ . <sup>c</sup> Calculated to fit $R_0 = 1.5$ , reflecting pre-vaccination estimate of MPVX $R_0$ in Ontario [17] via [18]. <sup>d</sup> Leaky-type. <sup>e</sup> Optimized parameter.

**Figure 1:**
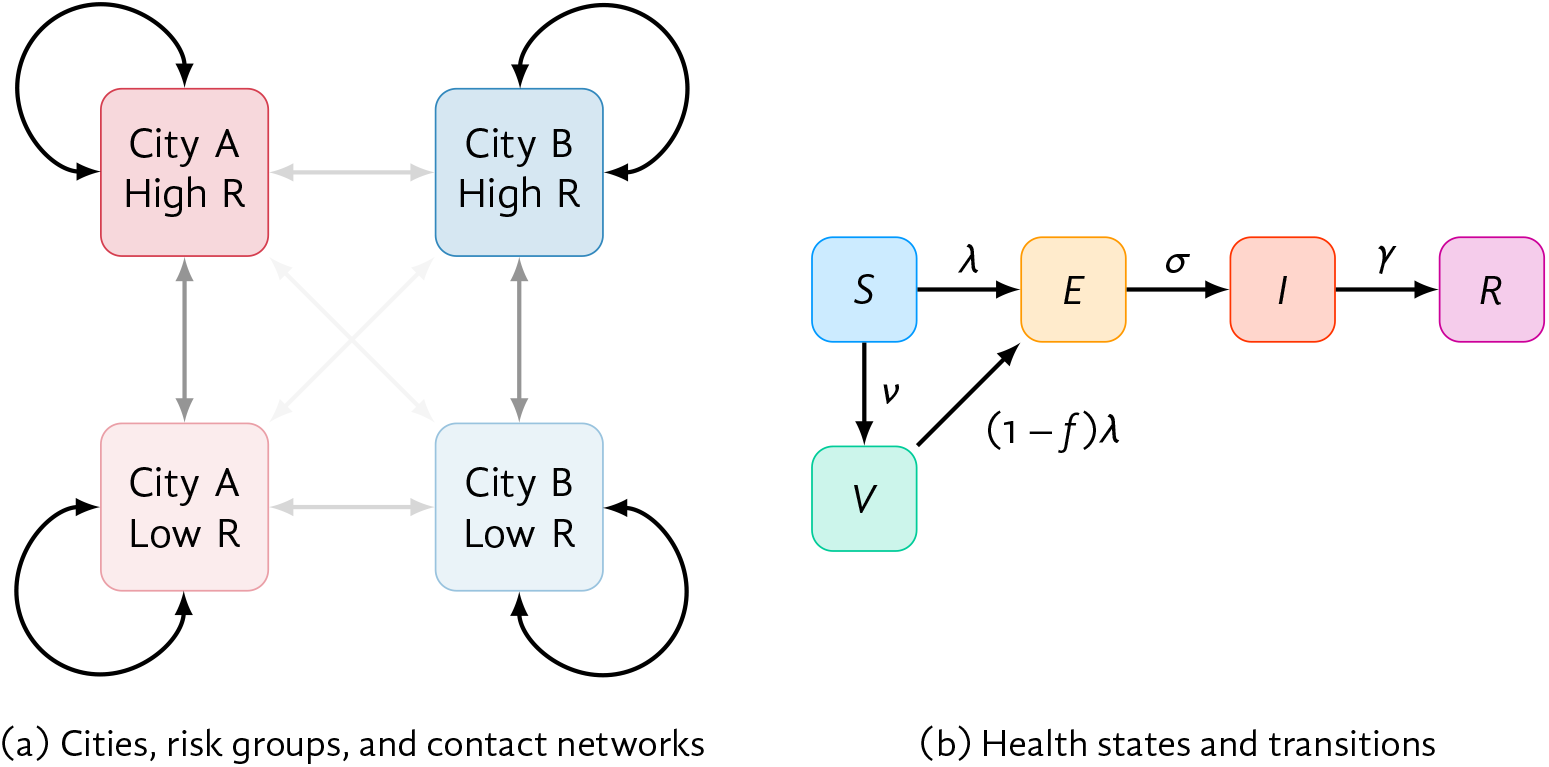
Model structure (a) High/Low R: risk groups; arrow opacity is proportional to contact network connectivity between groups. (b) *S*: susceptible; *V*: vaccinated; *E*: exposed; *I*: infectious; *R*: recovered. See Table A.1 and Appendix A for rate definitions.

We initialized all model runs with 10 imported/seed cases, distributed across the exposed and infectious stages proportionally by mean stage duration. We then simulated distribution of 5000 vaccine doses over 15 days, 60 days after initial cases were imported (though not necessarily detected). Doses were imperfectly prioritized to the higher risk group with 90% sensitivity — i.e. 4500 doses reach the higher risk group and 500 each the low risk group, reflecting early risk-based eligibility criteria in some jurisdictions [3].

Using this model, we explored optimal vaccine allocation between cities A and B over a range of epidemic conditions. For a given set of conditions, we defined the optimal vaccine allocation as that which resulted in the fewest cumulative infections by day 120 in both cities.^1^

We chose this 60-day time horizon and fixed 5000 vaccine doses to reflect a plausible medium-term optimization problem relevant to the early mpvx situation in Ontario. In reality, multiple changing time horizons may require consideration, different numbers of doses may become available, and different rates of vaccination may be possible. We aimed to obtain generalizable insights about the relationships between specific epidemic conditions and efficient geographic prioritization of vaccines during an outbreak.

As one specific example setting, we chose parameters representative of Toronto (city A) and another medium-sized Ontario city (city B), with gbmsm population sizes of 80,000 and 20,000, respectively, and 10% sexual/social network connectivity (*ϵ*_*c*_ = 0.9) [7]. We also modelled *R*_0_ = 2.0 in Toronto versus 1.5 in city B, reflecting differences in sexual network density as suggested by differential prevalence of bacterial sexually transmitted infections across Ontario cities [19,20]. Finally, we simulated 100% imported/seed cases in Toronto, reflecting early mpvx case distribution in Ontario [17]. We then compared two strategies of vaccine allocation by city: (a) proportional to population size; and (b) “optimal” (fewest infections by day 120).

Next, we performed a “grid sweep” of the following epidemic conditions, and identified the optimal vaccine allocation between cities A and B for each combination of conditions:

- relative size of city A versus B (1/4 to 4 times)
- relative epidemic potential in city A (*R*_0_ in city A from 1 to 2, versus fixed 1.5 in city B),^2^ adjusted via the sexual activity of the higher risk group in the city A
- between-city mixing (0 to 30% of all contacts formed randomly between cities)
- fraction of imported/seed cases in city A versus B (0–100%)

## 3 Results

Figure 2 illustrates modelled monkeypox incidence and cumulative infections in “Toronto” versus city B under different vaccine allocation strategies. Due to the larger population size, greater epidemic potential (*R*_0_), and having all imported/seed cases in Toronto in this scenario, allocating all 5000 vaccine doses to Toronto yielded the fewest infections by day 120: 1630 (c). Allocating vaccines proportionally to city size (b) yielded 1956 infections, while no vaccination (a) yielded 3466 infections.

**Figure 2:**
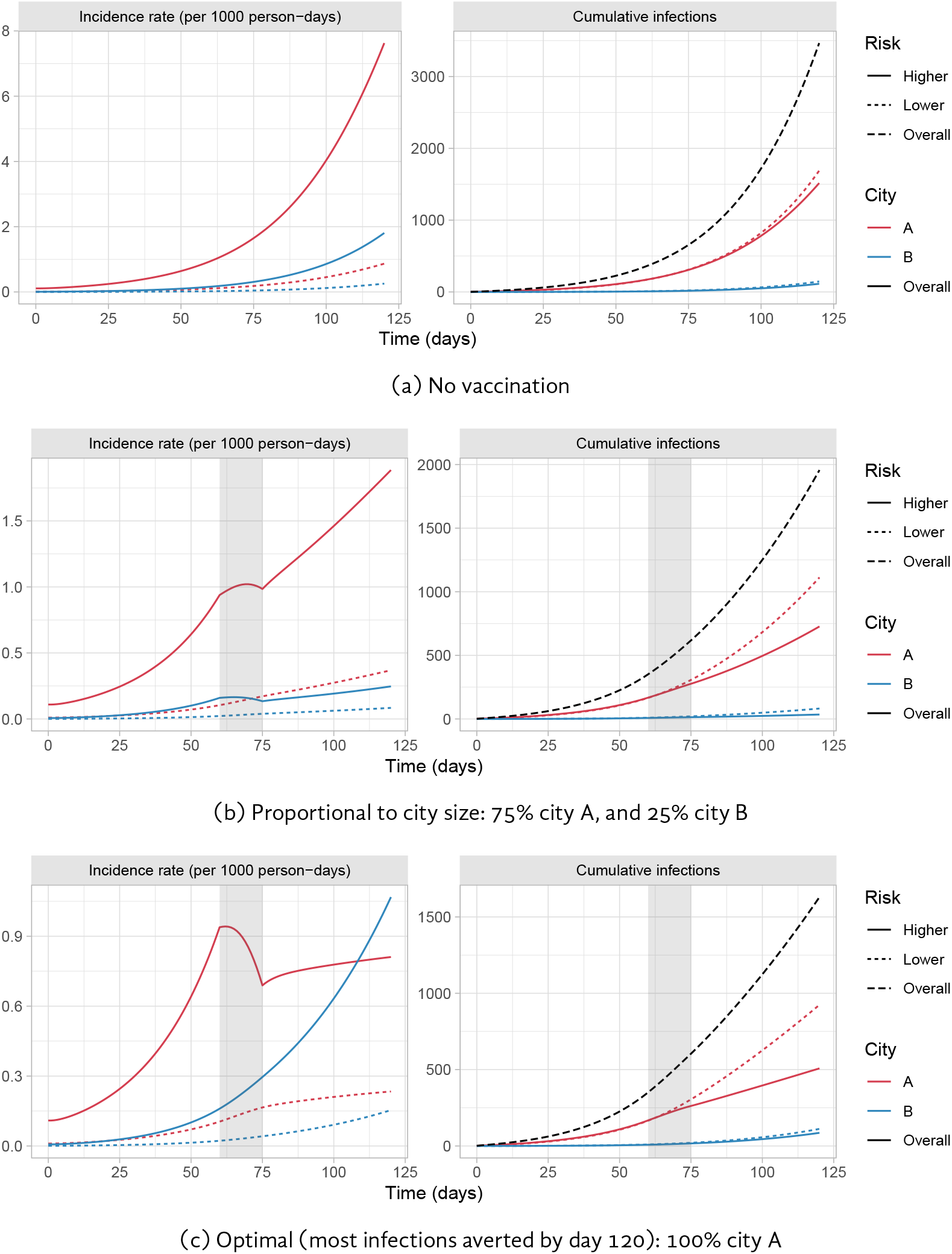
Modelled monkeypox incidence and cumulative infections in two cities under two different vaccine allocation scenarios Gray bar indicates period of vaccine roll-out (days 60–75). Cities loosely reflect Toronto and another medium-sized Ontario city.

As shown in Figure 2c, allocating most/all doses to one city (A) allows incidence to rise exponentially in the other city (B). However, this approach can still avert more infections overall over shorter time horizons, after which more doses may become available. Figure B.1 illustrates the opposite case (default model parameters in Table 1): two identical cities with equal seeding, where the optimal allocation is, unsurprisingly, equal between cities.

Figure 3 illustrates optimal vaccine allocation between cities A and B across different epidemic conditions. Figures B.2–B.5 further illustrate the absolute and relative numbers of infections averted under optimal allocation versus no vaccination (B.2–B.3), and versus vaccine allocation proportional to city size (B.4–B.5). Thus, Figures B.2–B.5 show under what conditions optimal allocation is most important.

**Figure 3:**
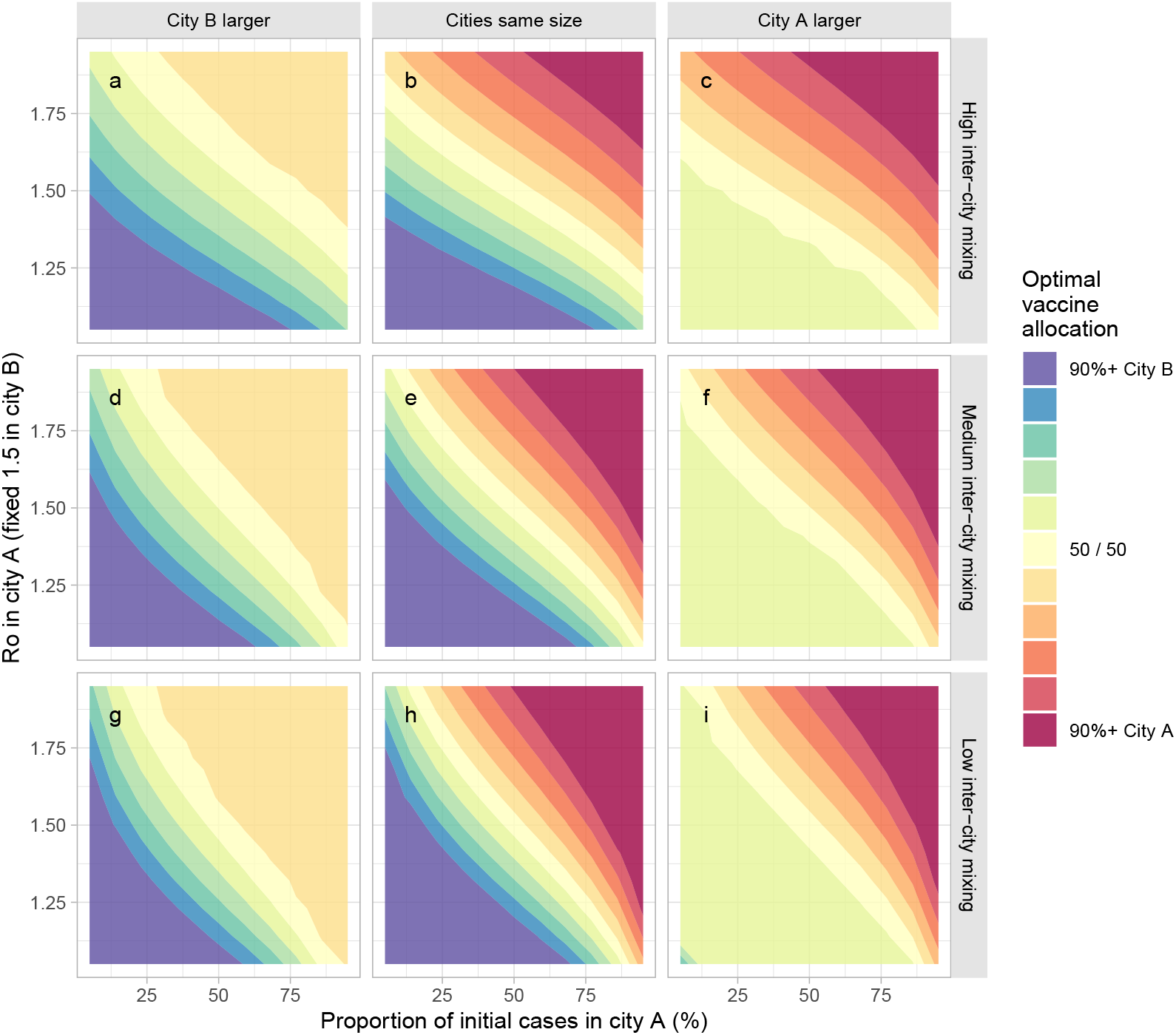
Optimal vaccine allocation between two cities under different epidemic conditions *R*_0_ in city A varies via the sexual activity among the high risk group in city A. Optimal allocation is defined as fewest cumulative infections by day 120. Larger city is 3 times the size of the other city. Most, moderate, and least inter-city mixing use *ϵ*_*c*_ = {0.8, 0.9, 0.95}, respectively.

The strongest determinants of optimal vaccine allocation were: relative epidemic potential (*R*_0_), share of seed cases, and city size; though city size was proportional to the size of the higher risk group under our modelling assumptions. Thus, if a larger city had large *R*_0_ and the majority of seed cases, it was best to allocate most/all doses to that city in our analysis (solid red/blue corners in Figure 3).

For smaller cities with large *R*_0_ and the majority of seed cases, it was sometimes possible to vaccinate the entire higher risk group; in this case, the remaining doses were best allocated to the higher risk group in the other city, yielding the plateaus (solid yellow triangles) in Figure 3: (a,d,g) upper right; (c,f,i) lower left. This plateau highlights how priority populations can change if/after high levels of coverage are achieved in other populations.

When cities with most/all seed cases had smaller *R*_0_, optimal allocation saw doses shared between cities (to varying degrees), suggesting that both risk-based (reflecting *R*_0_) and proximity-based (reflecting initial cases) prioritization strategies worked together to minimize transmission. In such cases, the other city necessarily had few/no seed cases but larger *R*_0_, to which the same findings apply. These conditions are represented by the yellow diagonal segments in all facets of Figure 3.

Finally, increased levels of mixing between cities mainly acted to reduce the influence of initial seed cases, and increase the influence of *R*_0_ on optimal allocation of vaccines to each city; this finding is visible in Figure 3 as stronger vertical gradients (contours are relatively more horizontal) in (a,b,c) with more inter-city mixing, versus stronger horizontal gradients (contours are relatively more vertical) in (g,h,i) with less inter-city mixing.

## 4 Interpretation

We sought to explore how different epidemic conditions could affect optimal allocation of a fixed supply of monkeypox virus (mpvx) vaccine across two weakly connected transmission networks (e.g. cities or jurisdictions). Under our modelling assumptions, we found that: vaccines could generally avert more infections when prioritized to a larger network, a network with more initial infections, and a network with greater epidemic potential (*R*_0_).

Although our study, for simplicity, focused on two weakly-connected networks, it highlights the importance of measuring outcomes for a population overall, by considering that geographies are comprised of interconnected networks. That is, while cities across Canada, and globally, feature important within- and between-city differences in size and configuration of transmission networks [21,22], and in access to interventions/services [20,23,24], ultimately these cities remain connected with respect to transmission, and cannot be considered in isolation over longer time horizons [7,22,25].

Within such interconnected settings, our findings are consistent with previous studies which show that prioritizing limited vaccine supply/resources to communities or settings with the highest epidemic potential (shaped by density and other features of the contact network) generally yields the greatest benefit for the population overall [4,5,26]. We also identified how key factors, such as number of imported cases and connections between networks, shape efficient early vaccine roll-out. While our model parameterization reflected gbmsm sexual networks in Ontario, our findings have wider implications for vaccine roll-out globally. The persistent absence of vaccine supply and roll-out in regions already endemic for mpvx outbreaks across West and Central Africa, including (although not yet reported) in the context of gbmsm and sexual minorities, poses the largest threat to the control and mitigation of mpvx globally [27], paralleling missed opportunities in achieving covid-19 vaccine equity [28].

Prioritizing based on risk also requires understanding risk. Early vaccine roll-out in Ontario reached Toronto, where cases were already detected, the population size was large, and rates of bacterial sexually transmitted infections suggested a potentially denser sexual network and thus, greater epidemic potential [13]. Our model implemented differential *R*_0_ between cities via contact rates; however, epidemic potential may also be linked to intervention access, including access to diagnoses and isolation support [23,29]. Thus, our findings signal the importance of characterizing the drivers of epidemic potential across jurisdictions and communities, including participatory, community-based surveillance and research into the contexts that lead to disproportionate risks at a network-level, not just an individual-level [30,31].

Our study aimed to provide fundamental and generalizable findings, and thus explored a broad sensitivity analysis to identify conditions that can shape optimal short-term vaccine allocation, with very limited supply. One limitation of our study is that we used a simple compartmental model, with only two risk groups; future work would benefit from more nuanced representations of risk, for example, using individual-based sexual network models. Second, our study only examined two transmission networks (“cities”); incorporation of additional networks could yield more interesting prioritization findings. However, we expect that the general principles and insights from two networks would apply across multiple networks.

## Data Availability

Monkeypox case series data were obtained from Public Health Ontario (see above).
All analysis code is available on GitHub (see below) from which it should be possible to reproduce all results in the paper.

https://github.com/mishra-lab/mpox-model-compartmental

## Acknowledgements

We thank: Kristy Yiu (Unity Health Toronto) for research coordination support; Huiting Ma, Linwei Wang, Oliver Gatalo, and Ekta Mishra (Unity Health Toronto) for support conceptualizing and parameterizing the model; and Mackenzie Hamilton (Unity Health Toronto) for her feedback on the manuscript. We also thank Toronto Public Health and members of the MPox Community Mobilization Group for their insights, ongoing engagement, and feedback on preliminary results.

## Contributions

JK and SM conceptualized and designed the study, and drafted the manuscript. JK developed the model, conducted the analyses, and generated the results. DT reviewed the results and contributed to manuscript writing.

## Data Availability

All analysis code is available at: github.com/mishra-lab/mpox-model-compartmental

### appendix

#### A Model Details

Table A.1 summarizes the notation used.

**Table A.1:** Notation

| Symbol | Definition |
| --- | --- |
| $c$ | city index $\in \{A, B\}$ |
| $r$ | risk group index $\in \{\text{high}, \text{low}\}$ |
| $y$ | type of contact index $\in \{\text{sexual}, \text{close non-sexual}\}$ |
| $N$ | population size |
| $C$ | contact rate |
| $Q$ | total contacts offered: $NC$ |
| $\epsilon$ | assortativity parameter $\in [1: \text{assortative}, 0: \text{random}]$ |
| $\lambda$ | incidence rate (force of infection) |
| $\beta$ | secondary attack rate<sup>a</sup> |
| $\sigma^{-1}$ | duration of latent/incubation period |
| $\gamma^{-1}$ | duration of infectious/symptom period |
| $\Phi$ | probability of contact formation |
| $\rho$ | proportion isolating among infectious |
| $\nu$ | vaccination rate |
| $f$ | vaccine effectiveness (leaky=type) |

All durations in days; all rates in per-day. <sup>a</sup> per-partnership transmission probability

##### A.1 Differential Equations

Equation (A.1) summarizes the system of differential equations for the SVEIR health states; each equation is repeated for each combination of city *c* (A, B) and risk group *r* (high, low) (4 total), but we omit the *cr* index notation for clarity.

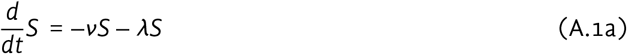

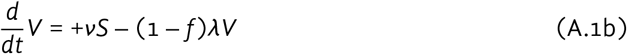

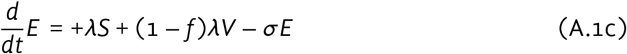

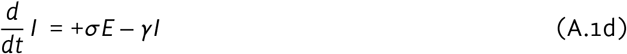

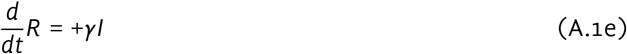

##### A.2 Incidence Rate

The incidence rate (force of infection) for non-vaccinated susceptible individuals in city *c* and risk group *r* (“group *cr*”) is defined as:

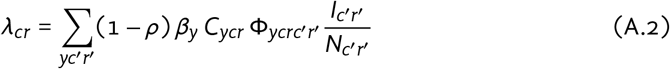

where: *ρ* is the proportion isolating among infectious; *β*_*y*_ is the transmission probability per type-*y* contact; *C*_*ycr*_ is the type-*y* contact rate among group *cr*; Φ_*ycrc*′*r*′_ is the probability of type-*y* contact formation with group *c*′*r*′ among group *cr*; and *N*_*cr*_ is the size of group *cr*.

Among vaccinated, the incidence rate is simply reduced by a factor (1 – *f*), where *f* is the vaccine effectiveness (leaky-type).

##### A.3 Mixing

Mixing between risk groups and cities was implemented using an adaptation of a common approach [1,2]. We denote the total contacts “offered” by group *cr* as: *Q*_*cr*_ = *N*_*cr*_*C*_*cr*_; and denote the margins *Q*_*c*_ = ∑_*r*_ *Q*_*cr*_; *Q*_*r*_ = ∑_*c*_ *Q*_*cr*_; and *Q* = ∑_*cr*_ *Q*_*cr*_. The probability of contact formation with group *c*′*r*′ among group *cr* is defined as:

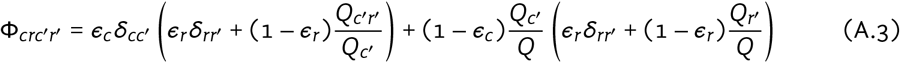

where: *δ*_*ii*′_ = {1 if *i* = *i*′; 0 if *i* * *i*′} is an identity matrix; and *ϵ*_*c*_, *ϵ*_*r*_ ∈ [0, 1] are assortativity parameters for mixing among cities and risk groups, respectively, such that *ϵ* = 1 yields complete group separation and *ϵ* = 0 yields completely random (proportionate) mixing. For clarity, we omit the index of contact type *y*, although *ϵ*_*r*_, *C*_*cr*_ and thus Φ_*crc*′*r*′_ are all further stratified by *y*.

##### A.4 City *R*_0_

The basic reproduction number *R*_0_ for each city was defined in the absence of vaccination and ignoring between-city mixing — i.e. with *ϵ*_*c*_ = 1. Following [3], we define *R*_0_ as the dominant eigenvalue of the city-specific next generation matrix *K*; matrix elements *K*_*rr*′_ are defined as:

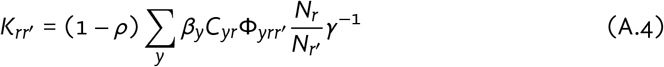

where: *ρ* is the proportion isolating among infectious; *β*_*y*_ is the transmission probability per type-*y* contact; *C*_*yr*_ is the type-*y* contact rate among group *r*; Φ_*yrr*′_ is the probability of type-*y* contact formation with group *r*′ among group *r*; *N*_*r*_ is the size of group *r*; and *γ*^−1^ is the duration of infectiousness.

##### A.5 Vaccine Allocation

Vaccination is modelled as distribution of 5000 doses over 15 days from day 60 (333 doses per day). Vaccines are prioritized to the high risk group with 90% sensitivity, such that 4500 doses actually reach the high risk group, and 500 doses are given to the lower risk group. Figure A.1 illustrates vaccination coverage/counts by city/risk group for an example allocation of 80% to city A and 20% to city B.

**Figure A.1:**
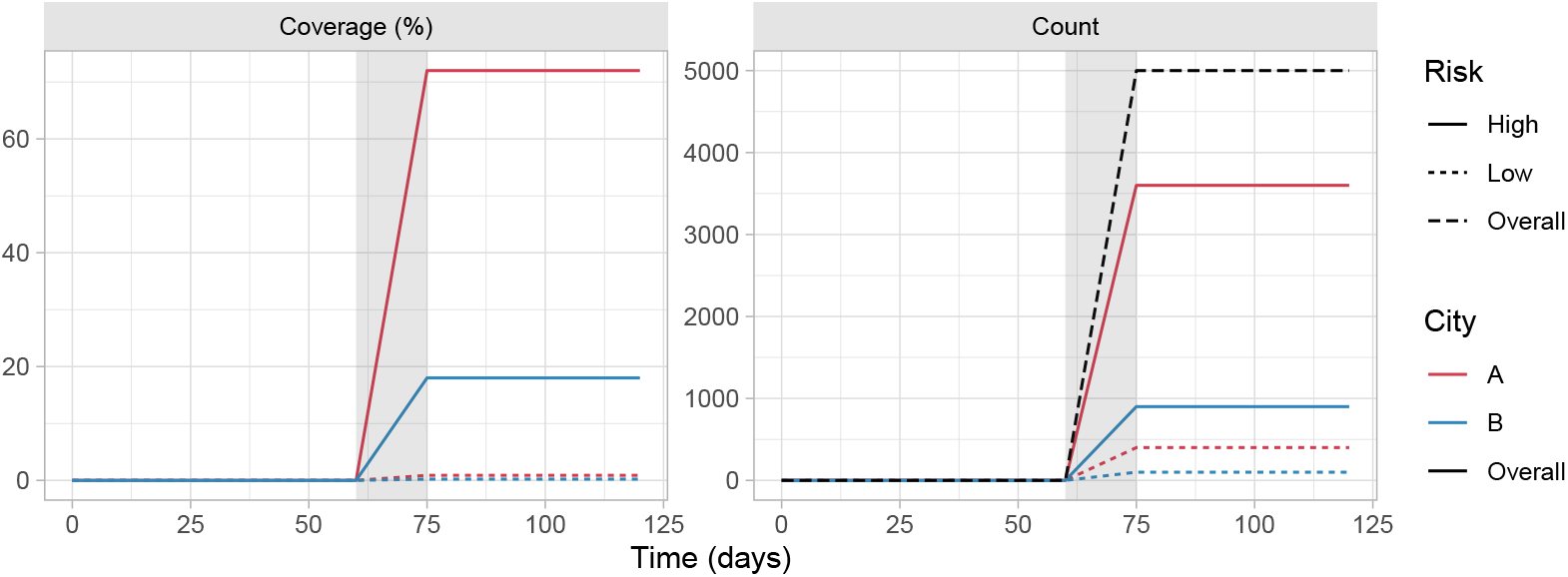
Example vaccine allocation: 80% to city A, and 90% to high risk group Gray bar indicates period of vaccine roll-out (days 60–75)

##### A.6 Parameterization

Model parameter values and stratifications are summarized in Table 1, repeated (verbatim) in Table A.2 for easier reference.

###### Sexual Behaviour

Parameterization of sexual behaviour was primarily informed by existing analyses conducted to support mathematical modelling of hiv-transmission among gbmsm in Canada [4, n.b. Appendix 3.2]. These analyses stratified gbmsm into 88–94% lower risk, with on average 4 sexual partners per-year (≈ .01 per day), and 6–12% higher risk, with approximately 6-times as many partners (≈ .07 per day). Our present model includes even greater partner numbers among the higher risk group (.10–.25 per day), partly to fit mpvx *R*_0_ ∈ [1, 2], and because the 6-fold value in [4] was mainly applied as a generalized proxy for 6-times higher HIV incidence. Weighted pooling of data from three studies [18–20] suggested that approximately 12% of respondents reported 20+ sexual partners in the past 6 months (≈ .11+ per day). Our mpvx model also models transmission risk per-partnership, versus per-contact (sex act) as in [4]; with high sar, mpvx transmission risk would be expected to be driven more by numbers of partners than by total contacts (sex acts).

###### Monkeypox Virus (mpvx)

Updated epidemiological data on mpvx infection and transmission in the context of the present epidemic are rapidly emerging [9,21]. In the absence of high-quality evidence on the secondary attack rate (sar) of sexual transmission, we assumed a relatively high sar of 0.9 (per-partnership), drawing on local patient histories, and in order to reproduce *R*_0_ ∈ [1, 2]. We estimated *R*_0_ ∈ [1, 2] using mpvx case data from Ontario [16] before widespread vaccine roll-out (2022 May 13 – July 4) using the EpiNow2 R package [17].

**Table A.2:** Model parameters, including default values and ranges explored via grid sweep

| Parameter | Stratum | Value | Range | Ref |
| --- | --- | --- | --- | --- |
| Population size | overall | 100,000 |  | [4]<sup>a</sup> |
|  | fraction in city A | .50 | [.20, .80] | <sup>a</sup> |
| Fraction higher risk | city A | .10 | [.01, .50]<sup>b</sup> | [4]<sup>a</sup> |
|  | city B | .10 |  | [4]<sup>a</sup> |
| Contact rate | close non-sexual, all | 1 |  | <sup>a</sup> |
|  | sexual, low risk | .01 |  | [4]<sup>a</sup> |
|  | sexual, high risk, city A | .178<sup>c</sup> | [.10, .25]<sup>b</sup> | [4,5]<sup>a</sup> |
|  | sexual, high risk, city B | .178<sup>c</sup> |  | [4,5]<sup>a</sup> |
| Assortativity | cities, all contacts | .90 | [.70, 1.0] | [6]<sup>a</sup> |
|  | risk, close non-sexual | 0 |  | <sup>a</sup> |
|  | risk, sexual | .50 |  | <sup>a</sup> |
| Per-contact SAR | close non-sexual | .05 |  | [7] |
|  | sexual | .90<sup>c</sup> |  | [5]<sup>a</sup> |
| Initial infections | overall | 10 |  | <sup>a</sup> |
|  | fraction in city A | .50 | [0.0, 1.0] | <sup>a</sup> |
| Duration of period | latent/incubation | 7 |  | [8–10] |
|  | infectious/symptoms | 21 |  | [9,11] |
| Fraction isolated among infected |  | .50 |  | [9]<sup>a</sup> |
| Vaccines available |  | 5000 |  | <sup>a</sup> |
| Vaccine effectiveness<sup>d</sup> |  | .85 |  | [12–14] |
| Vaccine prioritization sensitivity | high risk | .90 |  | [15]<sup>a</sup> |
| Vaccine allocation | city A | .50 | [0.0, 1.0]<sup>e</sup> | — |

All durations in days; all rates in per-day. SAR: secondary attack rate. <sup>a</sup> Assumed / representative. <sup>b</sup> Calculated to fit $R_0 \in [1, 2]$ . <sup>c</sup> Calculated to fit $R_0 = 1.5$ , reflecting pre-vaccination estimate of MPVX $R_0$ in Ontario [16] via [17]. <sup>d</sup> Leaky-type. <sup>e</sup> Optimized parameter.

In another model [5], the modelled *R*_0_ for a gbmsm sexual network was greater, even for smaller sar. Two main factors may explain this discrepancy in modelled *R*_0_ vs sar in [5] vs our model. First, isolation was not explicitly modelled in [5]; thus the reported sar in [5] can be considered as after considering isolation, i.e., reduced. Second, the branching process model in [5] captured greater risk heterogeneity than our model, and focused especially on capturing the highest levels of risk (“heavy tail”). Such heterogeneity is directly related to *R*_0_ through the coefficient of variation in contact rates [22]. Thus, this difference in model structure could further explain why modelled *R*_0_ would be greater in [5], for even similar sar. Finally, our aim was to obtain generalizable insights about network-level vaccine prioritization, rather than to model specific contexts within Ontario; as such, we do not expect our main findings to change with moderate changes to the model simplifications regarding transmission.

#### B Supplemental Results

Figure B.1 illustrates incidence rate and cumulative infections (similar results to Figure 2), for two cities identical in: size, *R*_0_, and imported/seed cases, under three vaccination scenarios: no vaccination, 100% allocation to city A, and equal allocation between cities. Equal allocation minimizes cumulative infections.

Figures B.2–B.5 illustrate cumulative infections averted by day 120 under “optimal” vaccine allocation: versus no vaccination (absolute: B.2, relative: B.3), and versus allocation proportional to city size (absolute: B.4, relative: B.5).

**Figure B.1:**
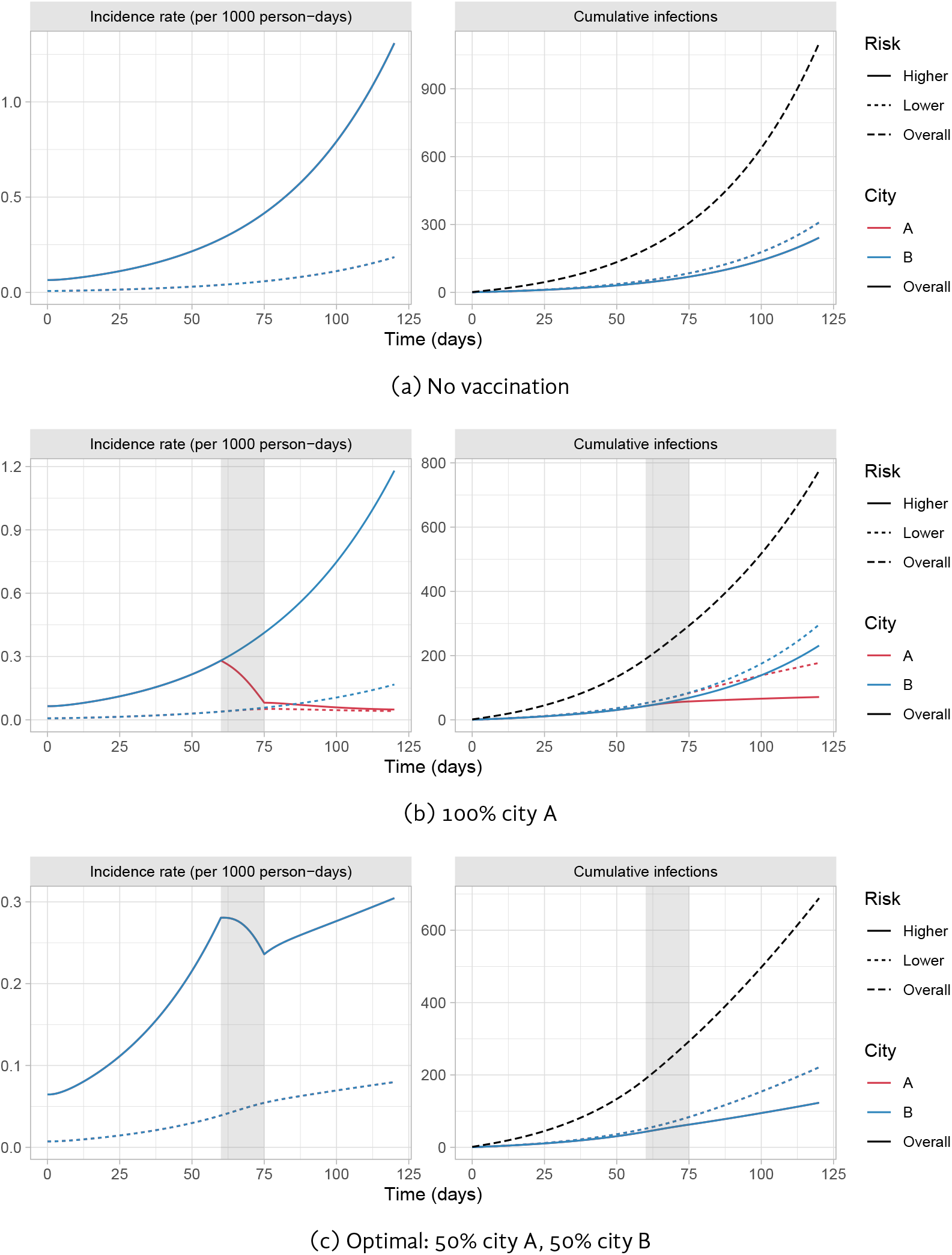
Modelled monkeypox incidence and cumulative infections in cities A and B with default parameters, under two different vaccine allocation scenarios Gray bar indicates period of vaccine roll-out (days 60–75).

**Figure B.2:**
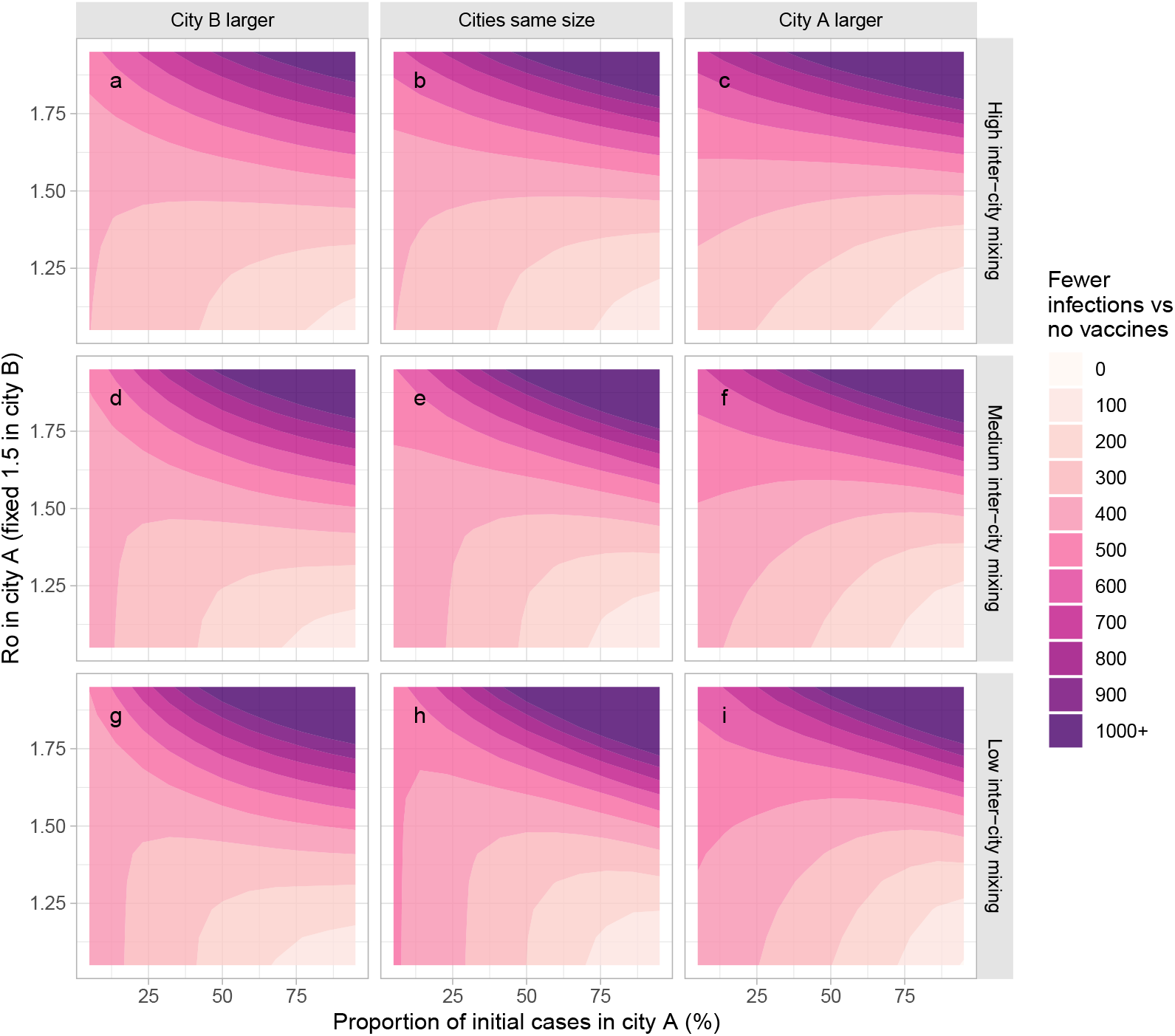
Absolute fewer infections under optimal vaccine allocation versus no vaccination *R*_0_ in city A varies via the sexual activity among the high risk group in city A. Optimal allocation is defined as fewest cumulative infections by day 120. Larger city is 3 times the size of the other city. Most, moderate, and least inter-city mixing use *ϵ*_*c*_ = {0.8, 0.9, 0.95}, respectively.

**Figure B.3:**
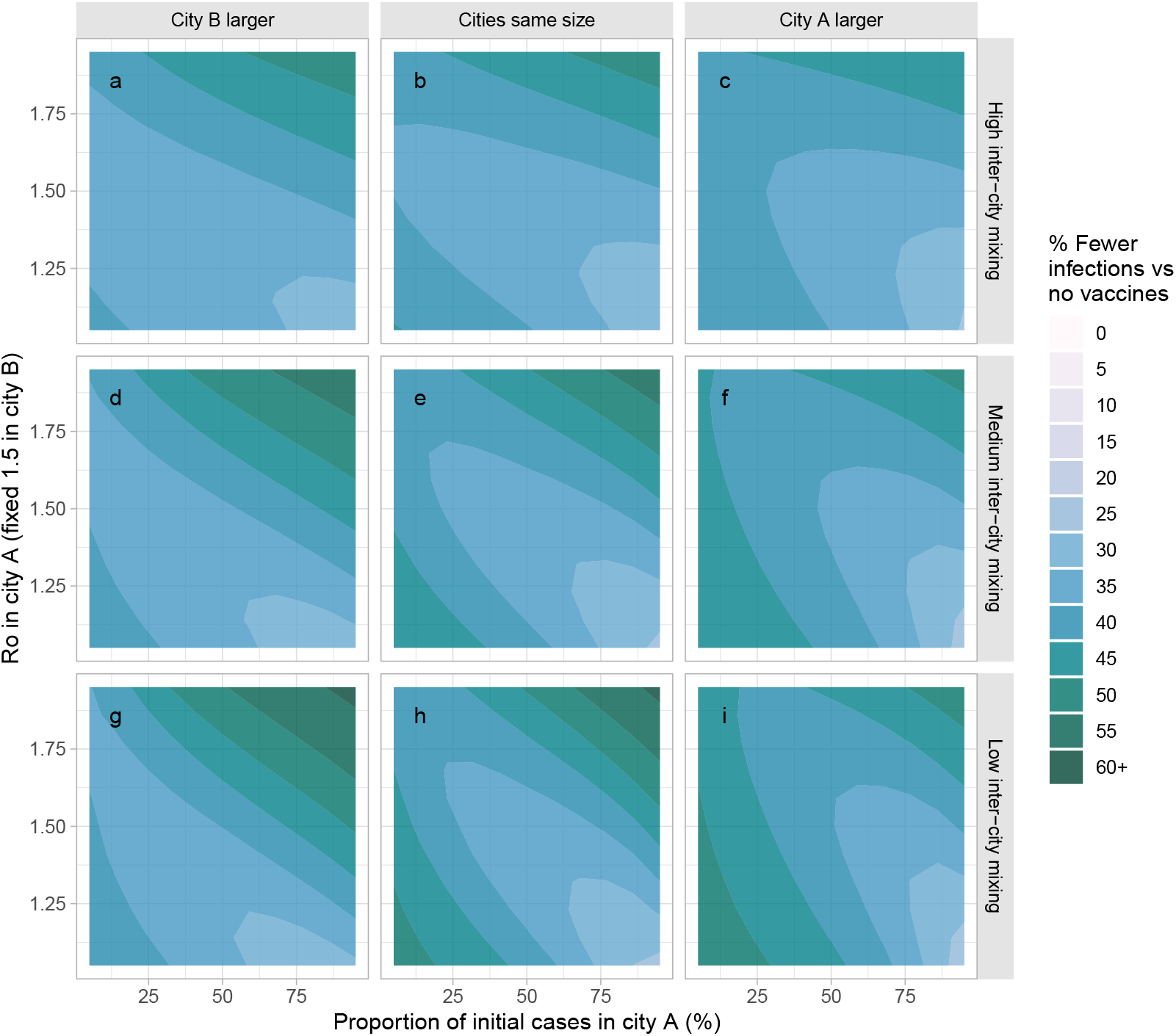
Relative fewer infections under optimal vaccine allocation versus no vaccination *R*_0_ in city A varies via the sexual activity among the high risk group in city A. Optimal allocation is defined as fewest cumulative infections by day 120. Larger city is 3 times the size of the other city. Most, moderate, and least inter-city mixing use *ϵ*_*c*_ = {0.8, 0.9, 0.95}, respectively.

**Figure B.4:**
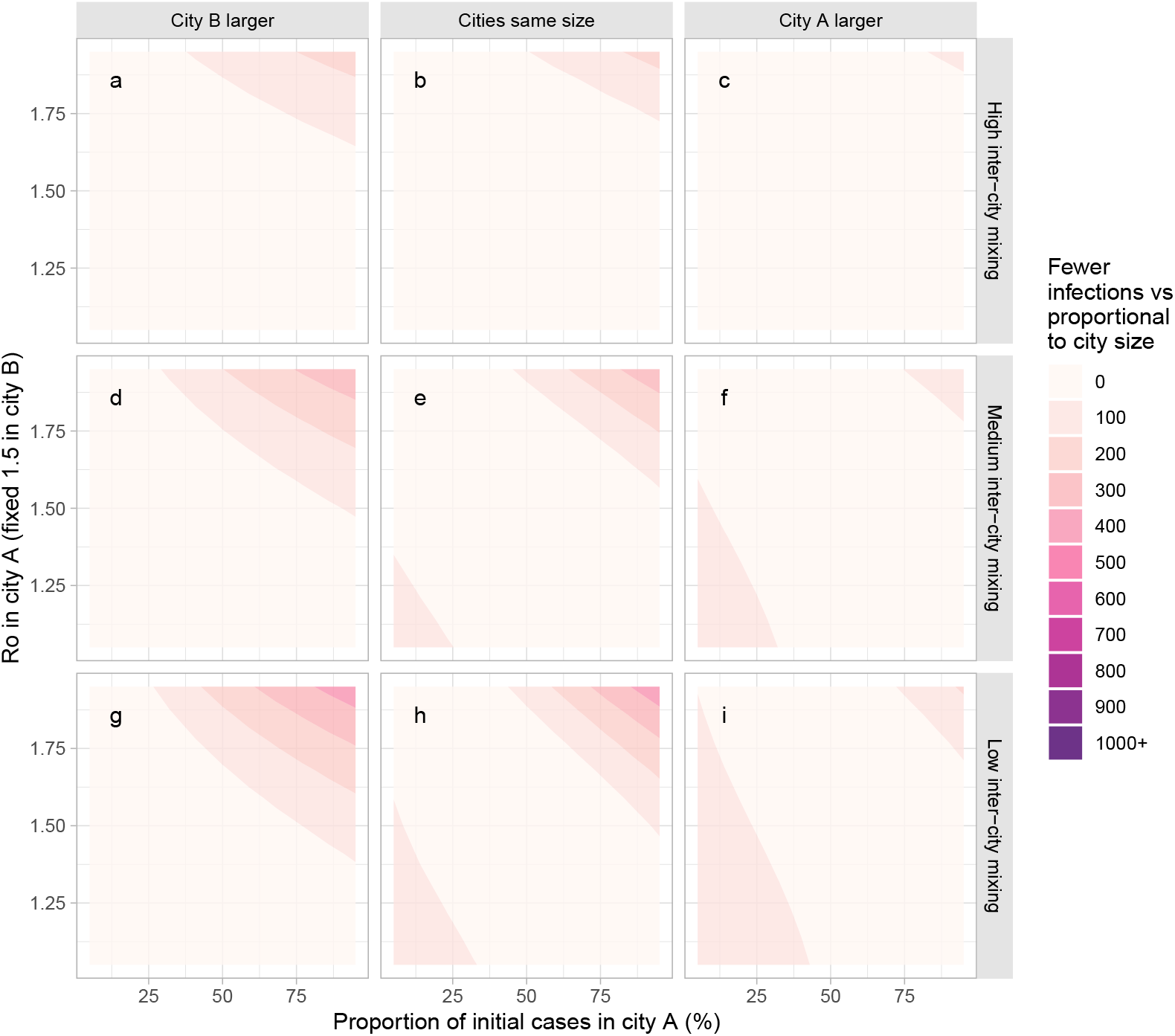
Absolute fewer infections under optimal vaccine allocation versus allocation proportional to city size *R*_0_ in city A varies via the sexual activity among the high risk group in city A. Optimal allocation is defined as fewest cumulative infections by day 120. Larger city is 3 times the size of the other city. Most, moderate, and least inter-city mixing use *ϵ*_*c*_ = {0.8, 0.9, 0.95}, respectively.

**Figure B.5:**
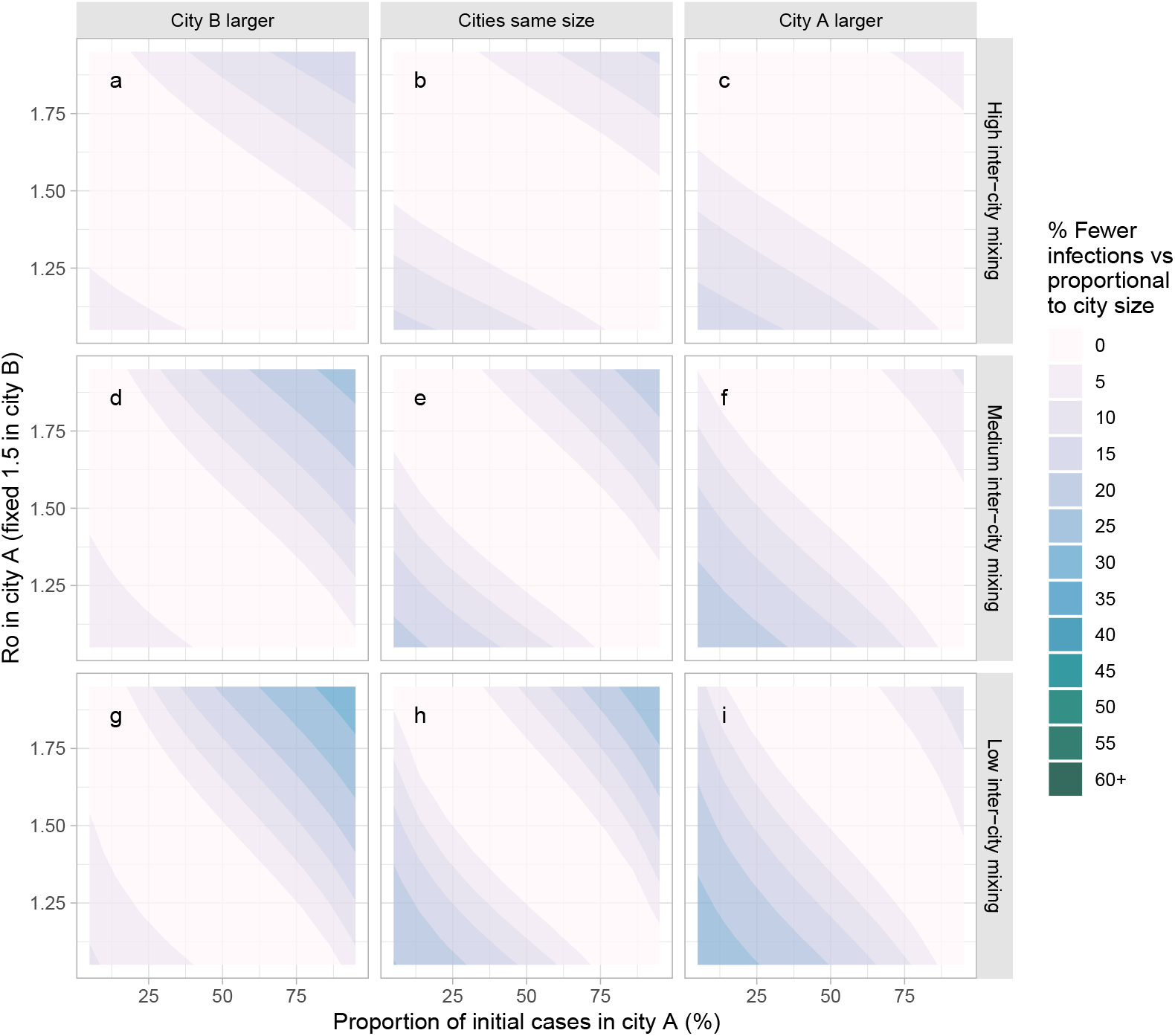
Relative fewer infections under optimal vaccine allocation versus allocation proportional to city size *R*_0_ in city A varies via the sexual activity among the high risk group in city A. Optimal allocation is defined as fewest cumulative infections by day 120. Larger city is 3 times the size of the other city. Most, moderate, and least inter-city mixing use *ϵ*_*c*_ = {0.8, 0.9, 0.95}, respectively.

## Footnotes

1 Optimal allocation was identified using the [optimize](https://www.rdocumentation.org/link/optimize?package=stats) function in R.

2 City-specific *R*_0_ calculated assuming no inter-city mixing.

